# Dose–response relationship between physical activity and mortality in adults with noncommunicable diseases: A systematic review and meta-analysis of cohort studies

**DOI:** 10.1101/2019.12.18.19014340

**Authors:** Wolfgang Geidl, Sabrina Schlesinger, Eriselda Mino, Lorena Miranda, Klaus Pfeifer

## Abstract

**Objective:** To investigate the relationship between post-diagnosis physical activity and mortality in patients with selected noncommunicable diseases, including breast cancer, lung cancer, type 2 diabetes (T2D), ischemic heart disease (IHD), chronic obstructive pulmonary disease (COPD), stroke, osteoarthritis, low back pain and major depressive disorders.

**Design:** Systematic review and dose–response meta-analysis.

**Data sources:** PubMed, Scopus and the Web of Science were searched systematically for English publications from the inception of the platforms until August 2018. Additionally, the search was updated in August 2019.

**Eligibility criteria:** Prospective observational studies examining the relationship between at least three physical activity categories and all-cause mortality or disease-specific mortality as the primary outcome.

**Results:** In total, 28 studies were included: 12 for breast cancer, 6 for T2D, 8 for IHD and 2 for COPD. The linear meta-analysis revealed that each 10 metabolic equivalent tasks (MET) h increase of physical activity per week was associated with a 22% lower mortality rate in breast cancer patients (Hazard Ratio [HR], 0.78; 95% CI: 0.71, 0.86), 12% in IHD patients (HR, 0.88; 95% CI: 0.83, 0.93), 30% in COPD patients (HR, 0.70; 95% CI: 0.45, 1.09) and 4% in T2D patients (HR, 0.96; 95% CI: 0.93, 0.99). The non-linear meta-analysis showed a regressive association with no threshold for the beneficial effect of physical activity on mortality.

**Conclusion:** Higher levels of post-diagnosis physical activity are associated with lower mortality rates in breast cancer, T2D, IHD and COPD patients, with evidence of a no-threshold and non-linear dose–response pattern.

**SUMMARY BOX:** *Existing findings:*

- Higher levels of physical activity are associated with a clear reduction in all-cause mortality in the general population.
- In the general population, the shape of the dose–response curve between levels of physical activity and reduced mortality rates is characterized by a regressive, non-linear effect.

*New findings:*

- Higher levels of post-diagnosis physical activity are associated with a clear reduction in all-cause mortality in adults with breast cancer, T2D, IHD and COPD.
- The shape of the indication-specific dose–response curves between post-diagnosis physical activity and mortality are characterized by a regressive, non-linear association with (1) no threshold for the beneficial effect, (2) pronounced reductions of mortality for lower levels of physical activity compared to those who are physically inactive and (3) no harmful effects at higher levels of physical activity.

## INTRODUCTION

Physical activity has been proposed as a form of treatment for people with noncommunicable diseases (NCDs).[1] There is evidence for the central role that physical activity plays in the health status of people with NCDs. Regular physical activity positively influences symptoms and comorbidities, physical fitness and health-related quality of life in more than 25 NCDs, including osteoarthritis, type 2 diabetes mellitus (T2D), stroke and clinical depression.[1] However, it is less clear whether higher levels of physical activity in adults with NCDs also reduce mortality rates and thus lead to longer life expectancies.

The current evidence for the general population regarding physical activity and mortality is comprehensive and unambiguous. Numerous large cohort studies have consistently demonstrated an inverse relationship between physical activity levels and mortality.[2] Meta-analyses with pooled data from these studies produce similar findings.[3, 4] Compared with the lower physical activity groups, the risk of premature death was remarkably reduced in the higher physical activity groups. The meta-analysis conducted by Samitz et al.[4] included 80 primary studies with a total of 1,338,143 participants and revealed that per one hour increment of moderate-intensity physical activity per week, the relative risk of mortality was reduced by 4%. In the updated physical activity guidelines for healthy adults from the U.S. Department of Health and Human Services,[5] a clear dose–response association between the volume of physical activity and mortality rates has been shown. The shape of the dose– response curve is characterized by a regressive, non-linear effect, where the greatest difference in mortality rates occurs among inactive and minimally active individuals. For higher physical activity levels, the dose–response curve flattens out. This means that the relative risk of mortality continues to decline with higher volumes of physical activity with no adverse effects on mortality, even at very high levels of physical activity.[5]

In adults with NCDs, the current evidence on dose–response relations between physical activity and mortality is considerably weaker and inconsistent. For T2D, the meta-analysis conducted by Kodama et al.[6] found that an increment of one MET (metabolic equivalent tasks) h/day of physical activity was associated with a 9.5% relative risk reduction in all-cause mortality, thereby suggesting that post-diagnosis physical activity levels may result in similar mortality risk reductions compared to the general population. In a meta-analysis for patients with cancer, comparably beneficial associations between physical activity and mortality rates were reported by Li et al.[7]. Moore et al.[8] pooled data from six cohort studies of 654,827 individuals and adjusted their analysis for several confounders, including preexisting NCDs. In contrast, they concluded that the longevity effects of physical activity vary according to the preexisting NCDs, with higher benefits of regular physical activity in terms of life expectancy for those with a history of cancer (7.0 y) and heart disease (6.2 y) compared to those without these diseases (3.7 y).[8] Current evidence from the US Physical Activity Guidelines Advisory Committee[9] reported a general relationship between higher post-diagnosis physical activity and lower mortality rates in five NCDs (breast or colorectal or prostate cancer, the cardiovascular condition of hypertension and T2D). However, the committee found few studies that have systematically quantified the dose–response relations between physical activity levels and mortality end-points in people with preexisting NCDs. Accordingly, their report concludes that dose–response relationships cannot yet be defined for adults with NCDs as a result of the limited information available.[9] Overall, it is unclear whether mortality rates in individuals with NCDs are affected by physical activity in the same way as mortality rates in the general population.

Thus, the objective of this study was to conduct a systematic review and dose–response meta-analysis of physical activity and mortality in people with selected NCDs. We aimed to define the dose–response relationship between post-diagnosis physical activity and mortality rates for nine NCDs with a high global burden of disease,[10] including low back pain, T2D, osteoarthritis, depressive disorder, chronic obstructive pulmonary disease (COPD), breast cancer, lung cancer, stroke and ischemic heart disease (IHD).

## METHODS

The method for this systematic review and meta-analysis was predefined in a published study protocol,[11] and registered at PROSPERO – the International Prospective Register of Systematic Reviews (registration number: CRD42018103357; available online at https://www.crd.york.ac.uk/prospero/display_record.php?RecordID=103357). This systematic review and meta-analysis is reported in compliance with the Preferred Reporting Items for Systematic Reviews and Meta-Analyses (PRISMA) Statement for Reporting Systematic Reviews and Meta-Analysis (see Supplementary File 1).[12]

### Search and data sources

A systematic search was conducted of PubMed, Scopus and the Web of Science from their inception to August 2018. This search was followed by a hand-search of the citations in the detected articles. The search was updated in August 2019 by using the forward citation search in Google Scholar for the articles that qualified for inclusion (see Supplementary File 2).

### Study selection

The eligibility criteria required the population to consist of adults with a physician-confirmed or self-reported diagnosis of one of the nine NCDs (osteoarthritis, low back pain, depressive disorder, IHD, T2D, stroke, COPD, lung cancer or breast cancer). Studies that investigated the association between physical activity and all-cause mortality as the primary outcome or any other indication-specific mortality as a primary or secondary outcome were included. For the dose–response meta-analysis, at least three categories of the exposure (i.e. physical activity) had to be reported in the original study. The eligible study design was that of a prospective observational nature. Non-English-language records, studies conducted on non-human subjects and duplicate data sets were not considered. No limit on publication year was imposed.

First, the literature identified through the electronic search was primarily assessed for eligibility by inspecting the titles and abstracts. We decided to divide the literature between three reviewers because of the large number of hits. Two additional reviewers were appointed to ensure the quality of the first screening process. In the second step, the full texts of the qualified studies were retrieved and critically evaluated for their final inclusion in the data collection process. The three reviewers independently assessed the articles for eligibility, and any discrepancies were resolved by discussions and when necessary, by adjudication from another reviewer.

### Data collection and items

The following details were extracted from the included publications: first author, year of publication, study name, design, country, mean follow-up time, total sample size, age, sex, mortality cases in total and per physical activity category, exposure categories, diagnosis and mortality ascertainment, relative risks and corresponding 95% CIs of the multivariate-adjusted models. Thirteen authors of the selected studies were contacted for additional data on physical activity. However the original data from two authors did not allow for an estimation of physical activity levels in MET-h/week (meaning that these studies were excluded), and two authors provided information on physical activity dosage.[13, 14]

### Risk of bias in individual studies

The Cochrane tool for assessing the “Risk Of Bias In Non-randomised Studies - of Interventions” (ROBINS-I) was used to estimate the risk of bias and endorse conclusions closer to the truth.[15] The tool includes seven domains that lead to the risk of bias. These domains are due to 1) confounding, 2) selection of participants, 3) exposure assessment, 4) misclassification during follow-up, 5) missing data, 6) measurement of the outcome, and 7) selective reporting of results. The included studies were independently evaluated by two assessors (EM, LM). Any inconsistencies in the evaluations were documented and then discussed with a third member of the research team (WG) and resolved by mutual agreement.

### Statistical analysis

The meta-analysis was performed using Stata statistical software (Version 15, StataCorp, College Station, TX, US). We pooled aggregated data using the random effects meta-regression model, as suggested by DerSimonian and Laird,[16] assuming random variance of the true effect of physical activity among studies, especially due to diversity in assessment methods. For studies that reported results from one cohort in stratified estimates (e.g. separately for men and women), a fixed effect model was used to combine the effects for the whole cohort and include it in the meta-analysis. We conducted the linear dose–response association between physical activity per 10 MET-h/week and all-cause mortality via the method used by Greenland and Longnecker and presented via forest plots.[17, 18] For this analysis, the number of cases and person-years, the quantification of the exposure and RRs with the corresponding 95% CIs of at least three categories were needed. If information was missing, the distributions of cases and person-years were estimated using the total number of cases and the total number of participants plus the follow-up period, as previously described.[19] If the lowest category was not used as a reference, the reported risk estimates were recalculated using Orsini et al.’s[20] method to ensure comparability. The data on the volume of physical activity were converted into a unit of MET-h/week. If a study reported the exposure categories as ranges, then for each category, the midpoint between the lower and upper limit was calculated. For open categories, we assumed that the width was the same as the adjacent category. A potential non-linear association was evaluated using a restricted cubic spline model with three knots at the 10th, 50th and 90th percentile of frequency of the exposure.[18] The indication of nonlinearity was tested using a likelihood ratio test.

The heterogeneity was described using the measure of inconsistency (I^2^), and tau^2^ was used to measure the variance between the included studies.[21] A subgroup analysis and meta-regression were performed to explore the heterogeneity across studies. The analyses were stratified by demographic variables (age, geographic area), follow-up duration (< 10 and ≥10 years), death cases (<100, 100–500 and ≥ 500), method of physical activity assessment (questionnaire and interview), risk of bias (moderate and serious) and additional disease-specific relevant factors (e.g. menopausal status in breast cancer). Publication bias was investigated through various visual and statistical tools, including funnel plots and Egger’s test for small-study effects, where asymmetry with a significance level of *p* < 0.1 suggests publication bias.[22, 23]

## RESULTS

The systematic database search yielded 44,518 publications in total. Three additional studies were identified from the reference lists. Full texts were retrieved and screened for 183 articles with the potential for inclusion. Twenty-eight studies satisfied the inclusion criteria for only four out of the nine NCDs; breast cancer (*n* = 12), T2D (*n* = 6), IHD (*n* = 8) and COPD (*n* = 2) (see Figure 1 for a detailed flow diagram).

**Figure 1.**
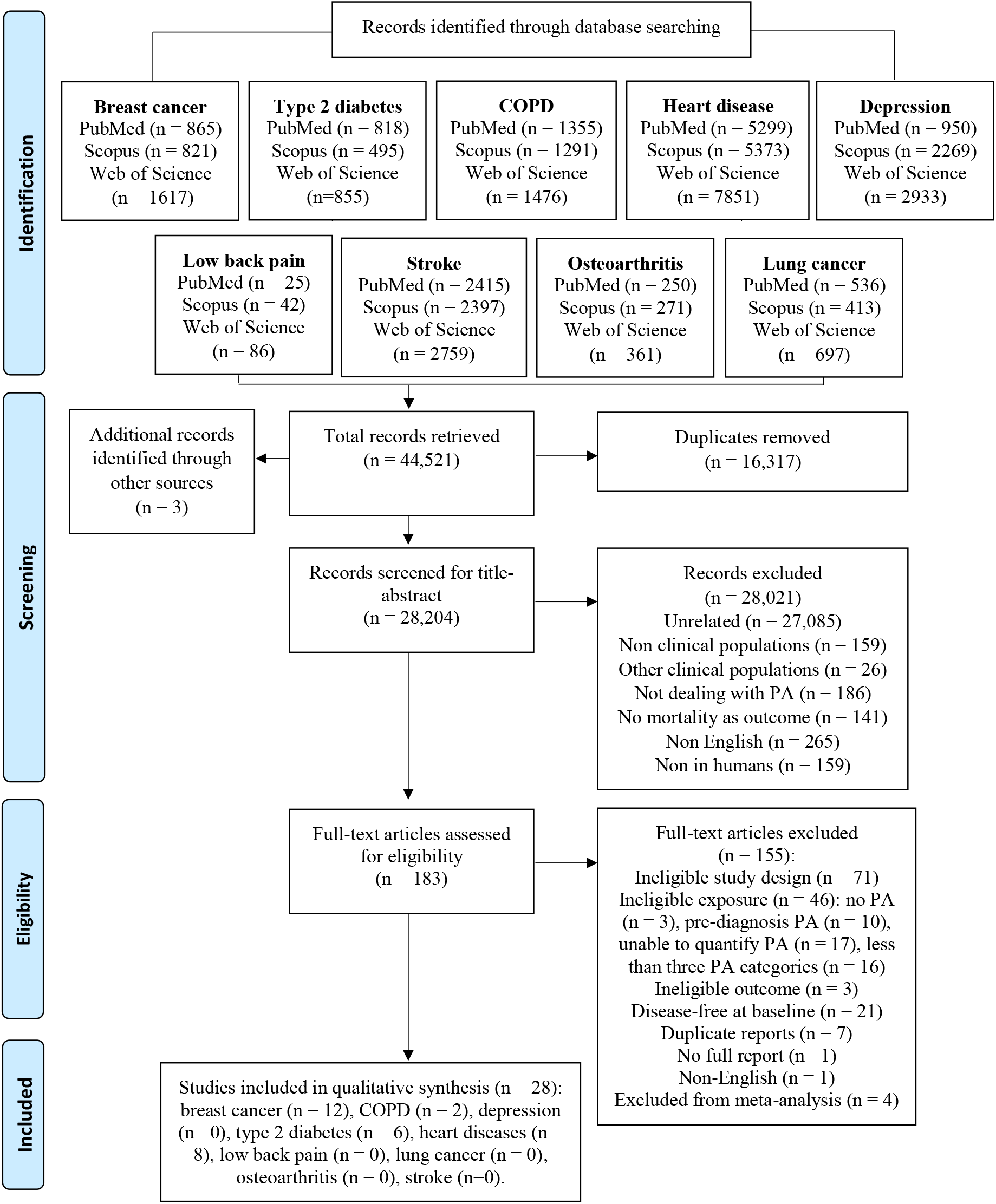
PRISMA flow diagram.

### Study characteristics

The 28 included studies were all published during the past two decades and based in numerous countries throughout the world. All the included studies had a prospective observational design. More specifically, there were 25 cohort studies that included two prospective cohort follow-ups to case-control studies as well as three follow-up studies of RCTs.[13, 24, 25] The sample sizes varied considerably from 435[26] to 15,645[27], with a total of 27,248 participants diagnosed with breast cancer, 32,221 with T2D, 4,784 with COPD and 42,027 with IHD.

The follow-up duration ranged from 3.3[28] years to 18.4 years.[29] A summary of the main characteristics of the cohorts is displayed in Table 1. All-cause mortality was reported as the primary outcome in all included studies. Other reported outcomes were breast cancer mortality, recurrence and new primary events, cardiovascular disease mortality, IHD mortality and respiratory mortality. All exposure assessments of physical activity were based on self-or interviewer-administered questionnaires. The time from diagnosis to physical activity measurement varied from three to six months post-diagnosis[30]. The longest follow-up was 14 years.[31] Detailed information on the measurement instruments for physical activity assessment can be found in Supplementary File 3. Exposure categories were presented as the volume of physical activity in MET-h/week[14, 24–28, 32–44] calorie expenditure,[45] duration of physical activity,[46] frequency of physical activity[47] and nominal categories.[13, 29–31, 48, 49]

**Table 1.**
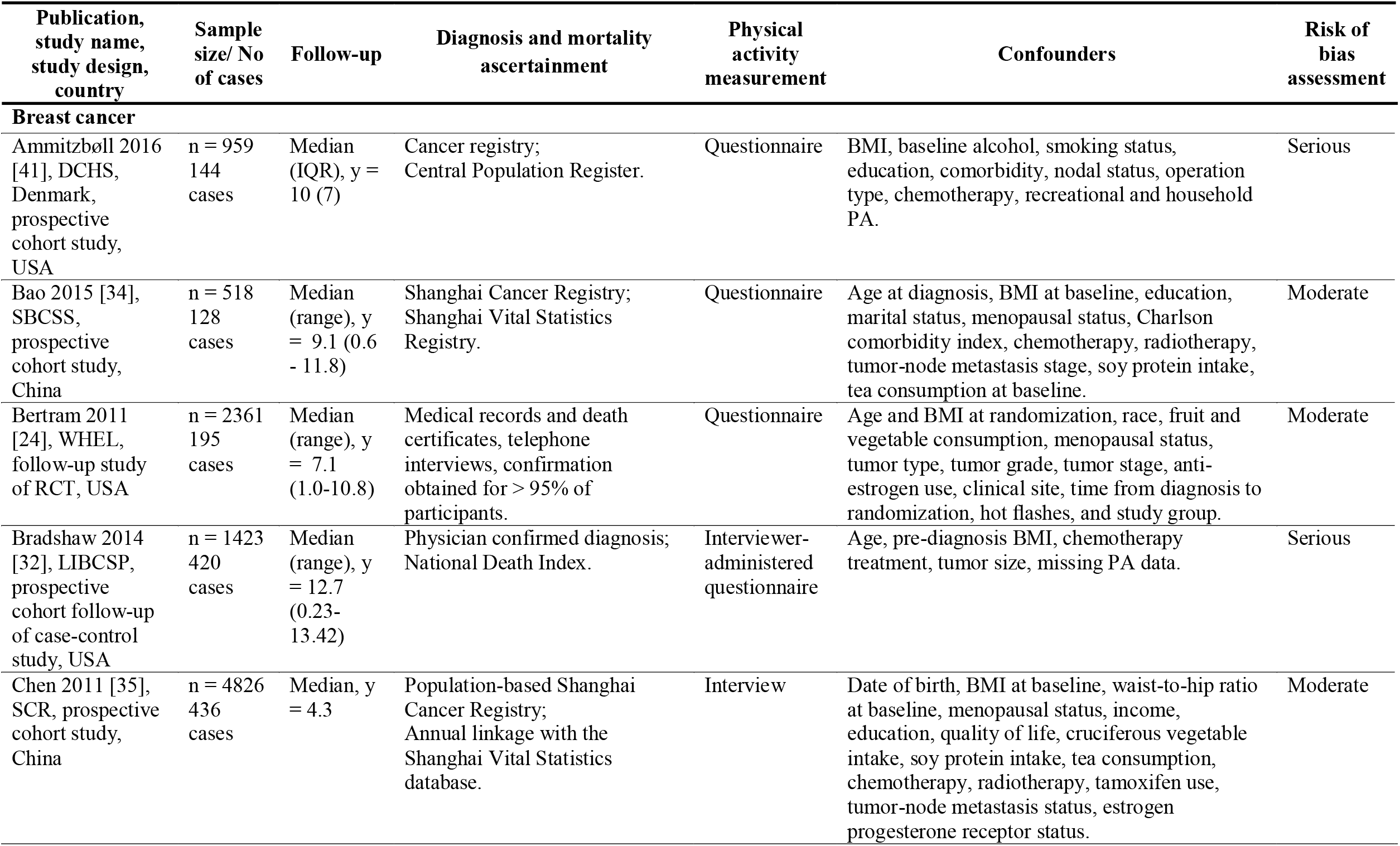
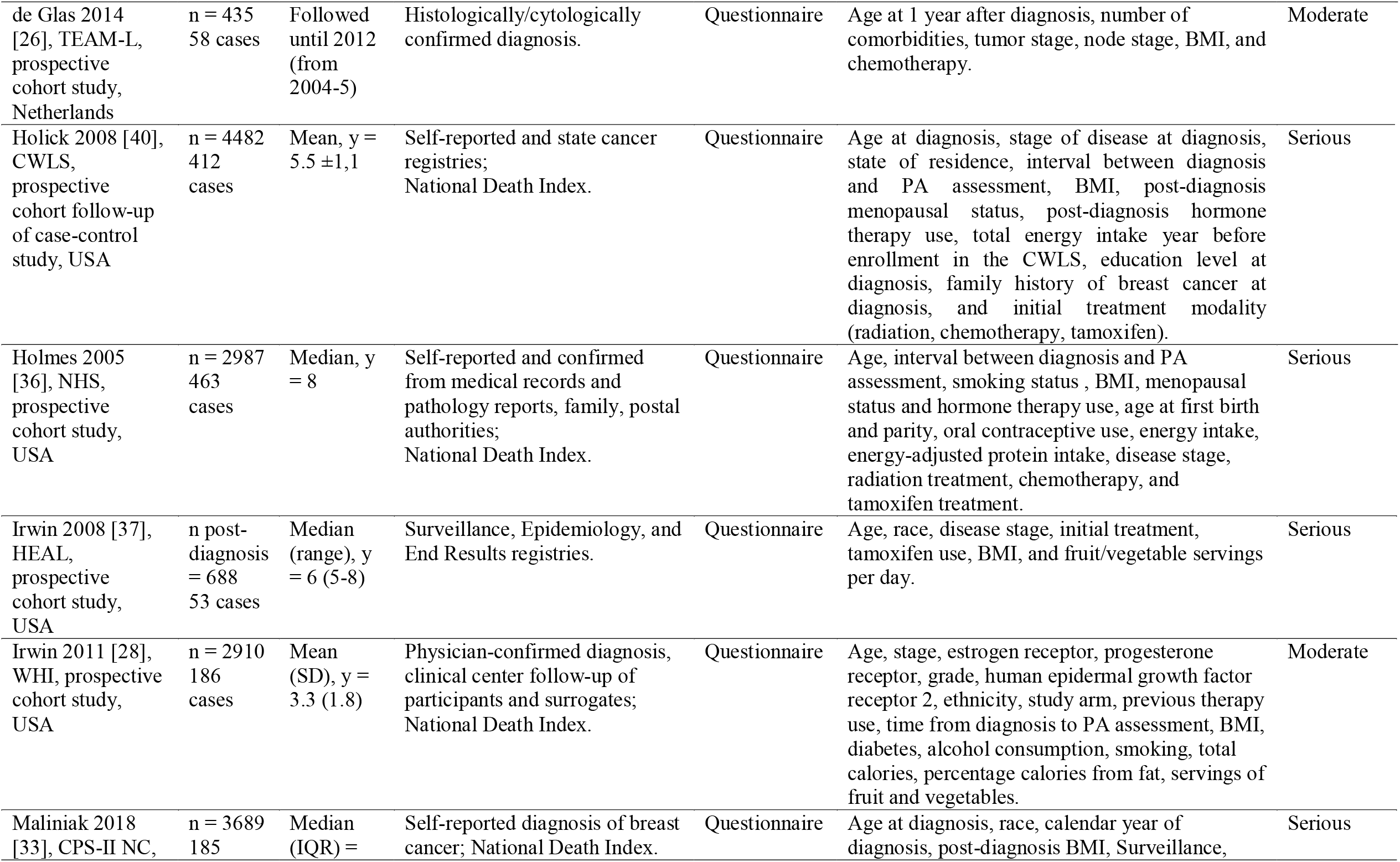
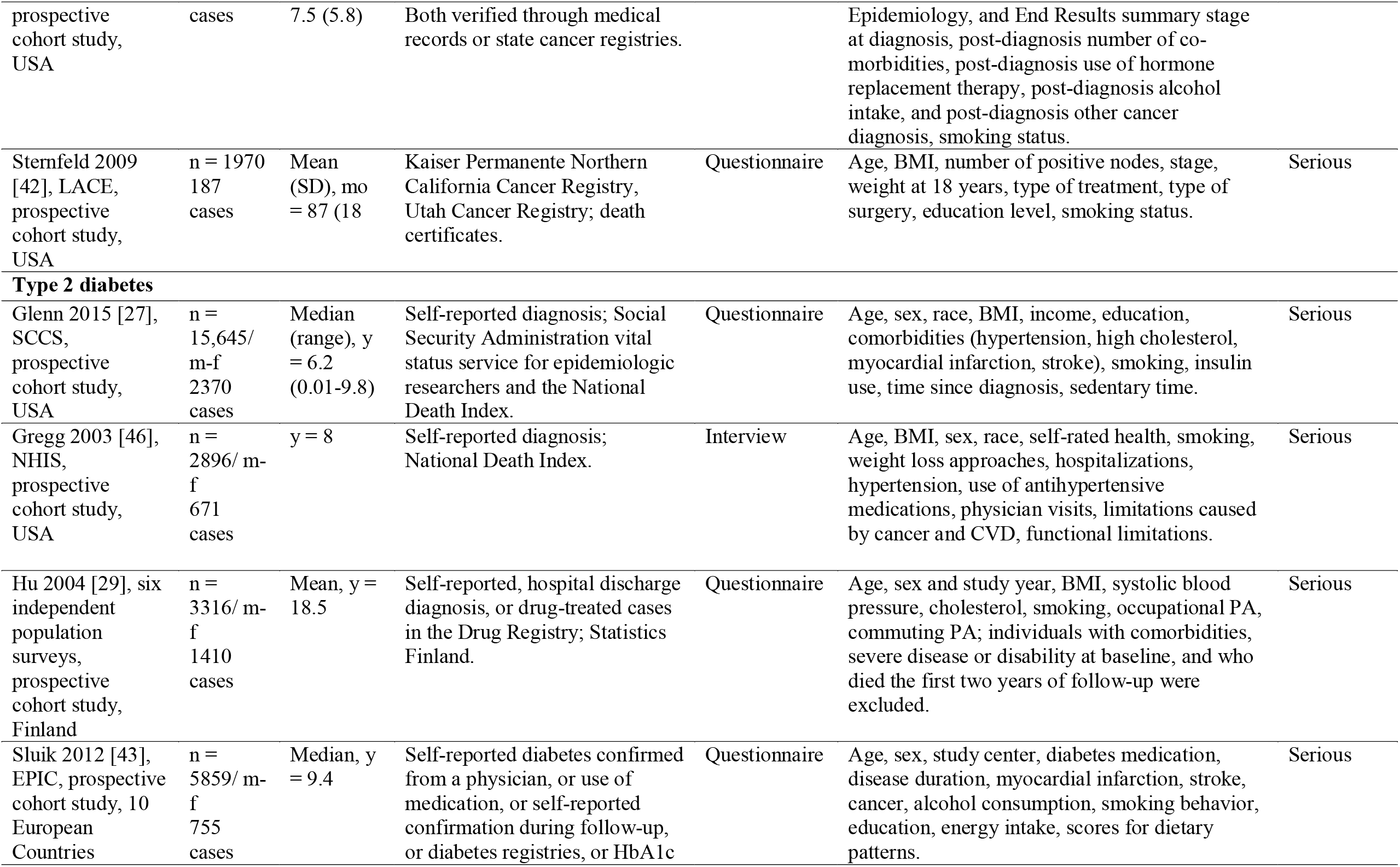
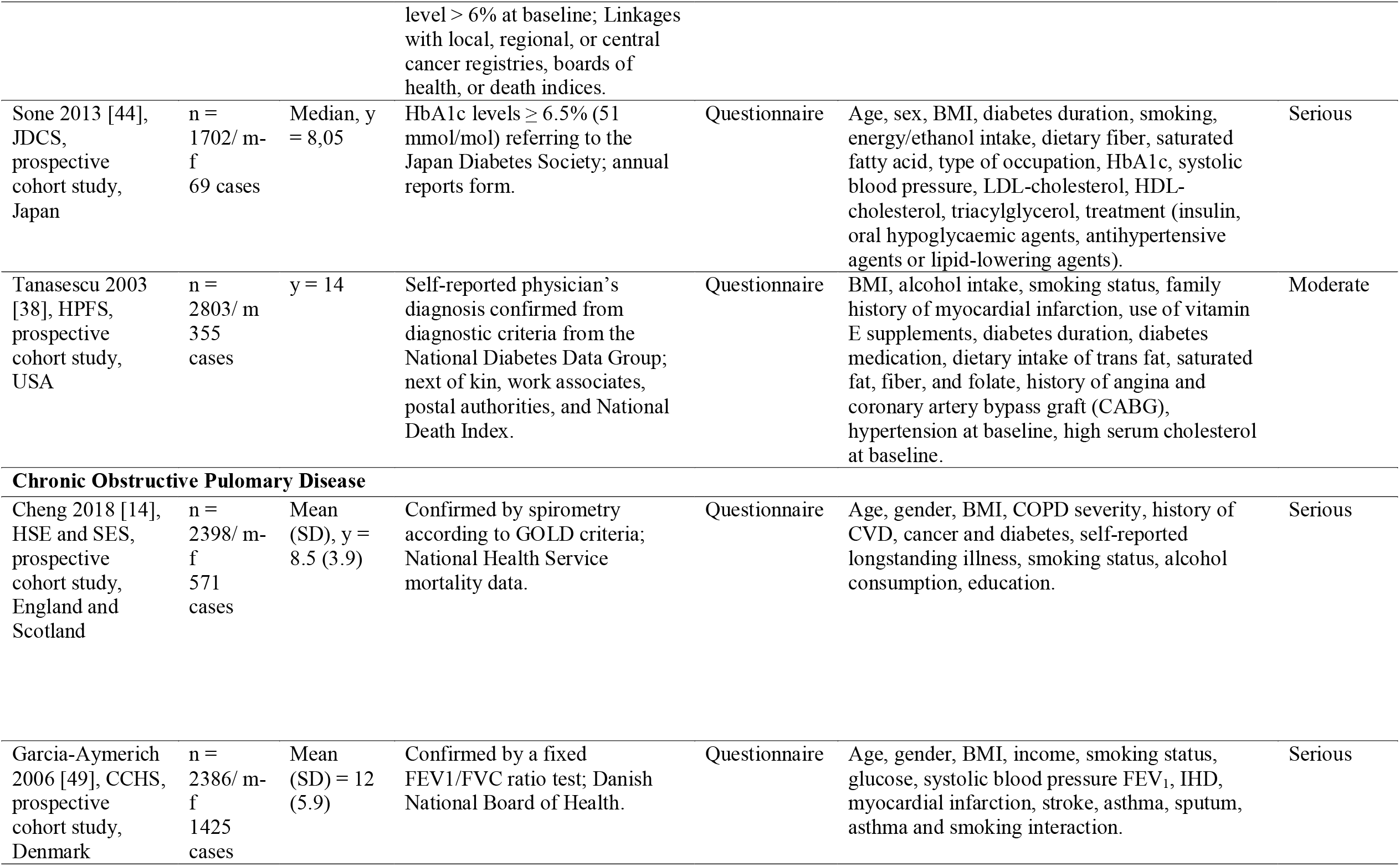
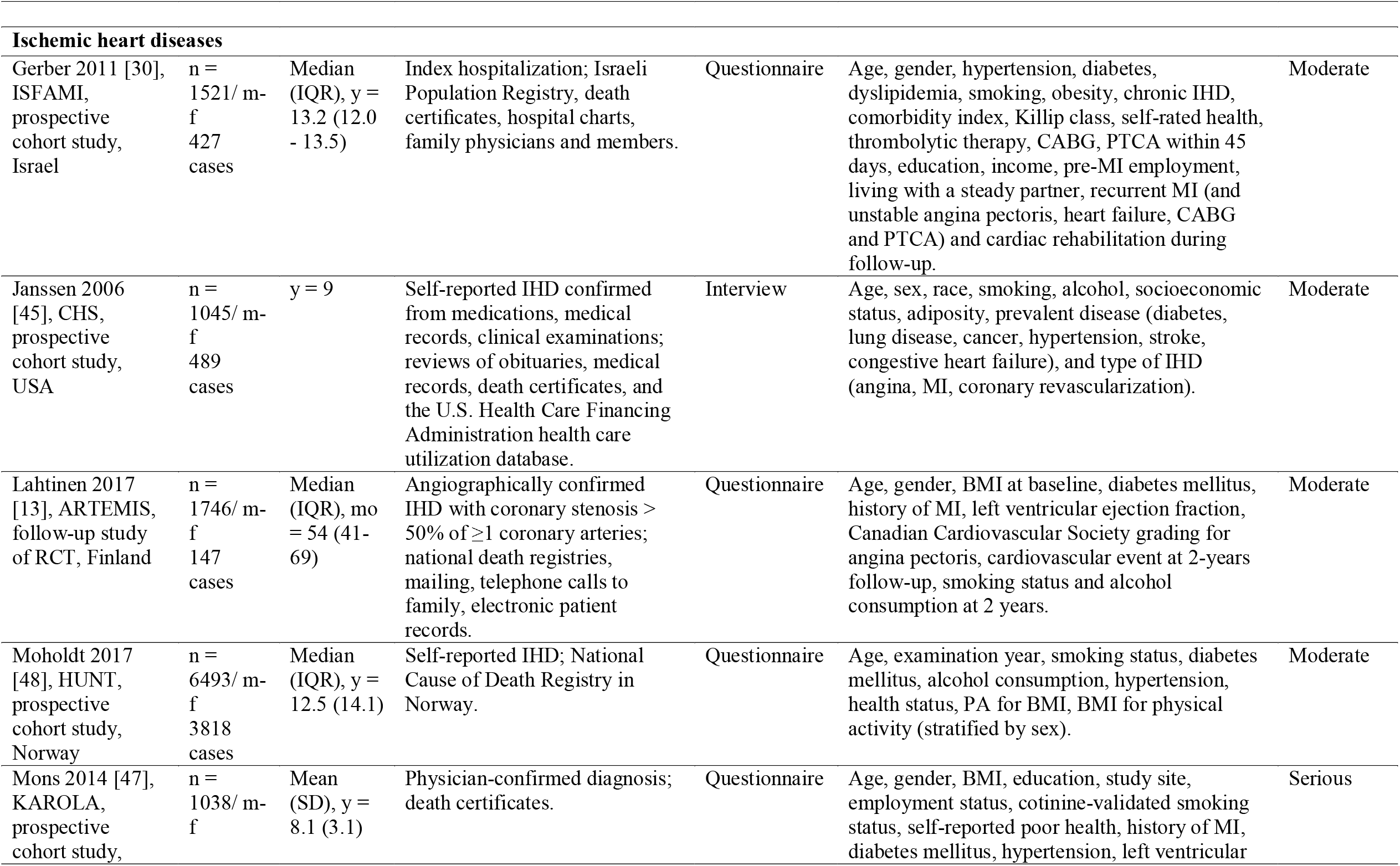
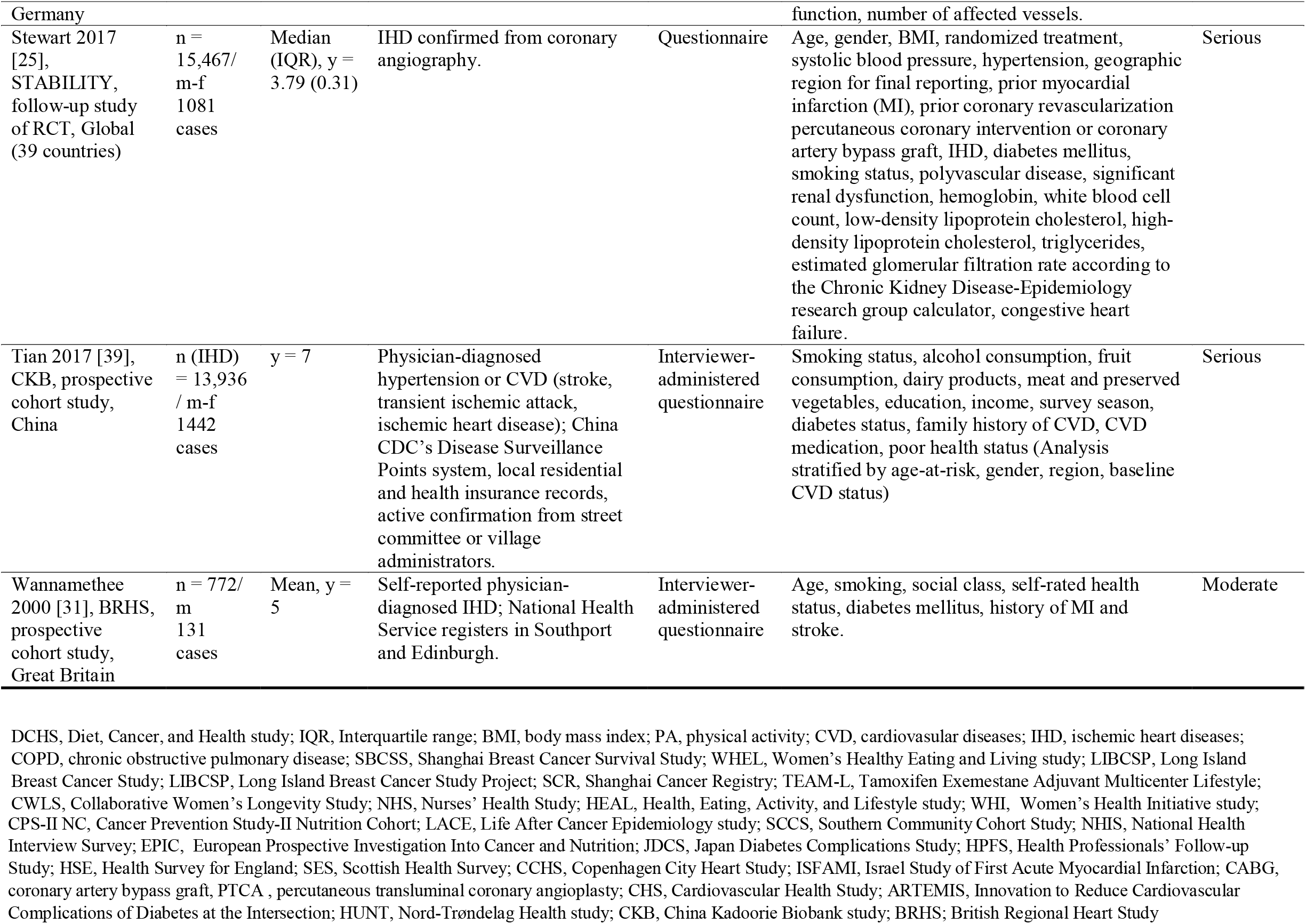
Main characteristics of the included studies.

### Risk of bias in included studies

In terms of the seven domains of ROBINS-I, no study had a low risk of bias. From the 28 publications assessed, 11 were evaluated to have a moderate risk of bias,[13, 24, 26, 28, 30, 31, 34, 35, 38, 45, 48] and the remaining studies had a serious risk of bias. The main domains that introduced bias were the confounding domain and the domain of deviations from intended interventions. It is important to note that due to the assessment of physical activity based on self-reports, a potential misclassification of physical activity could not be excluded. Therefore, 82% of the studies were rated with an unknown risk of bias in the domain of the classification of physical activity. The risk of bias for each domain in the 28 studies is shown in Figure 2. In addition, Table 1 contains the final risk of bias evaluation across the studies, and Supplementary File 4 includes the detailed results of the risk of bias assessment.

**Figure 2.**
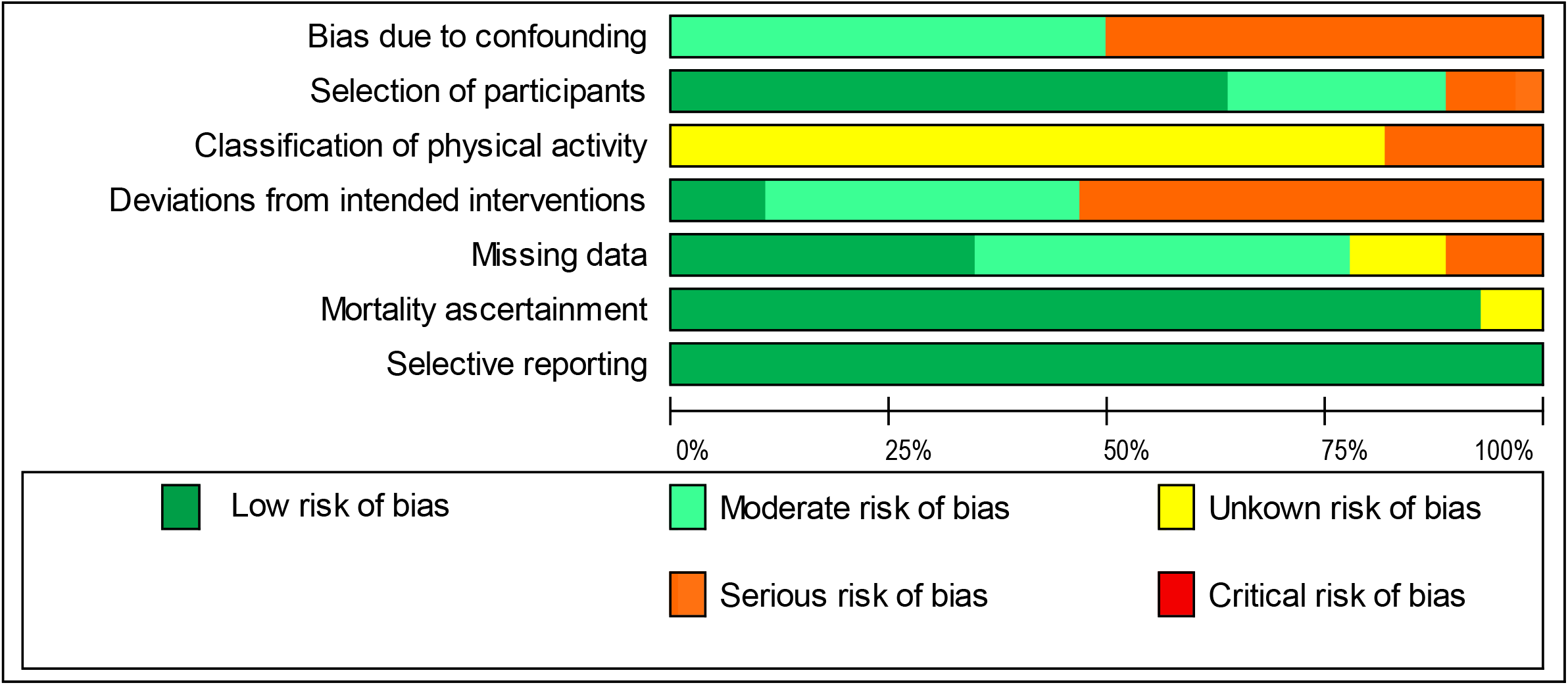
Risk of bias graph.

### Post-diagnosis physical activity and all-cause mortality

We examined the relationship between post-diagnosis physical activity and all-cause mortality in breast cancer, T2D, IHD and COPD populations. The results of the linear dose– response meta-analyses are presented in Figure 3. Physical activity was associated with lower mortality rates in persons with breast cancer, T2D, COPD and IHD. For every 10 MET-h increase of physical activity per week, the summary hazard ratio (SHR) decreased by 22% in people with breast cancer (HR, 0.78; 95% CI: 0.71, 0.86), by 12 % in people with IHD (HR, 0.88; 95% CI: 0.83, 0.93) and by 30% in people with COPD (HR, 0.70; 95% CI: 0.45, 1.09). The mortality rates in people with T2D reduced by 4% for every 10 MET-h/week (HR, 0.96; 95% CI: 0.93, 0.99). Ten MET-hours/week is equivalent to 180 minutes of walking or 86 minutes of running.[50]

**Figure 3.**
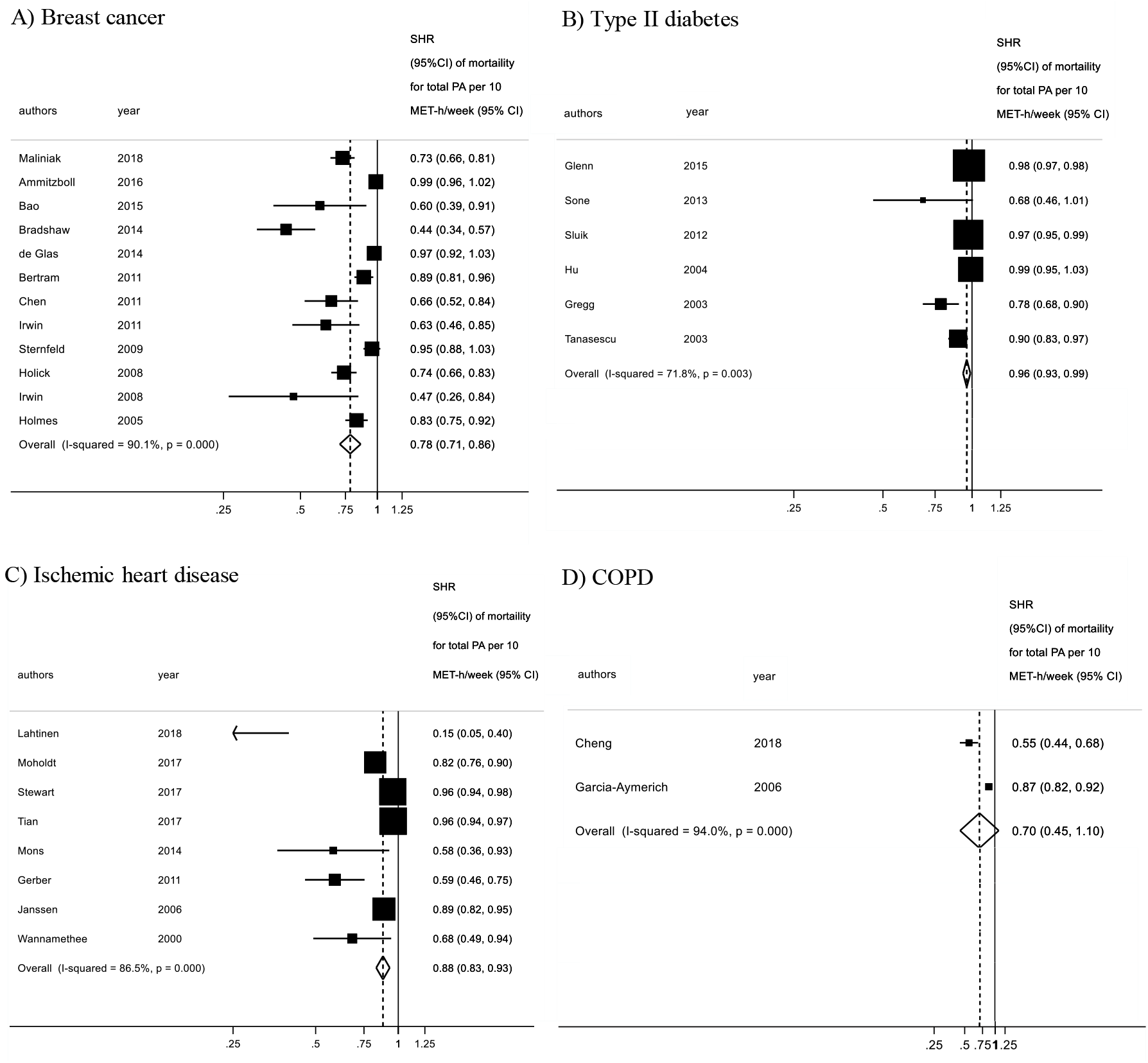
Linear dose–response meta-analysis for the association between post-diagnosis physical activity and all-cause mortality.

There was evidence of high heterogeneity between the included studies for all the target groups, specifically breast cancer (I^2^ = 90.1%), T2D (I^2^ = 72.7 %), IHD (I^2^ = 86.5%) and COPD (I^2^ = 94.0%). The subgroup analysis for breast cancer (Supplementary File 6, Table S6.1) highlighted that the subgroup difference is statistically significant (*p* = 0.018) for the follow-up variable only, meaning that the length of follow up can modify the observed associations between physical activity and mortality. None of the other included variables (age, geographic area, death cases, method of physical activity assessment, risk of bias, menopausal status in breast cancer) explained the amount of between-study variance (*p* > 0.05).

The funnel plot for breast cancer as well as T2D did not suggest the presence of publication bias, and Egger’s test confirmed that there was no apparent evidence of bias. However, the funnel plot for IHD studies was asymmetrical, and the test for small-study effects evidenced that publication bias could be present (*p* < 0.001) (Supplementary File 5). This should be interpreted with caution, however, as only eight studies were included in the funnel plot, and the statistical tests for publication bias are criticised for having low power.[51, 52] Furthermore, it should be noted that studies with smaller sample sizes[13, 30, 31, 47] have reported higher beneficial effects of physical activity. Thus, the detected asymmetry could be related to high heterogeneity.

Figure 4 presents the non-linear dose–response meta-analysis among the four NCD populations. The results for breast cancer (*n* = 12), T2D (*n* = 6), IHD (*n* = 8) as well as COPD (*n* = 2) indicated a non-linear dose–response relationship between post-diagnosis physical activity presented in MET-h/week and all-cause mortality (*p* _for non-linearity_ < 0.001). The curves for breast cancer, T2D and IHD show the steepest drop between 0 MET-h/week and 20 MET-h/week; the COPD curve drops more markedly between 0 and 10 MET-h/week. After this, the curves flatten out. For diabetes and COPD, significantly higher physical activity levels (up to around 90 MET-h/week) are also associated with further positive effects on mortality rates. For breast cancer, there appears a plateau with no additional effects on mortality with more than 45 MET-h/week. For COPD, the curve can only be calculated up to 30 MET-h/week.

**Figure 4.**
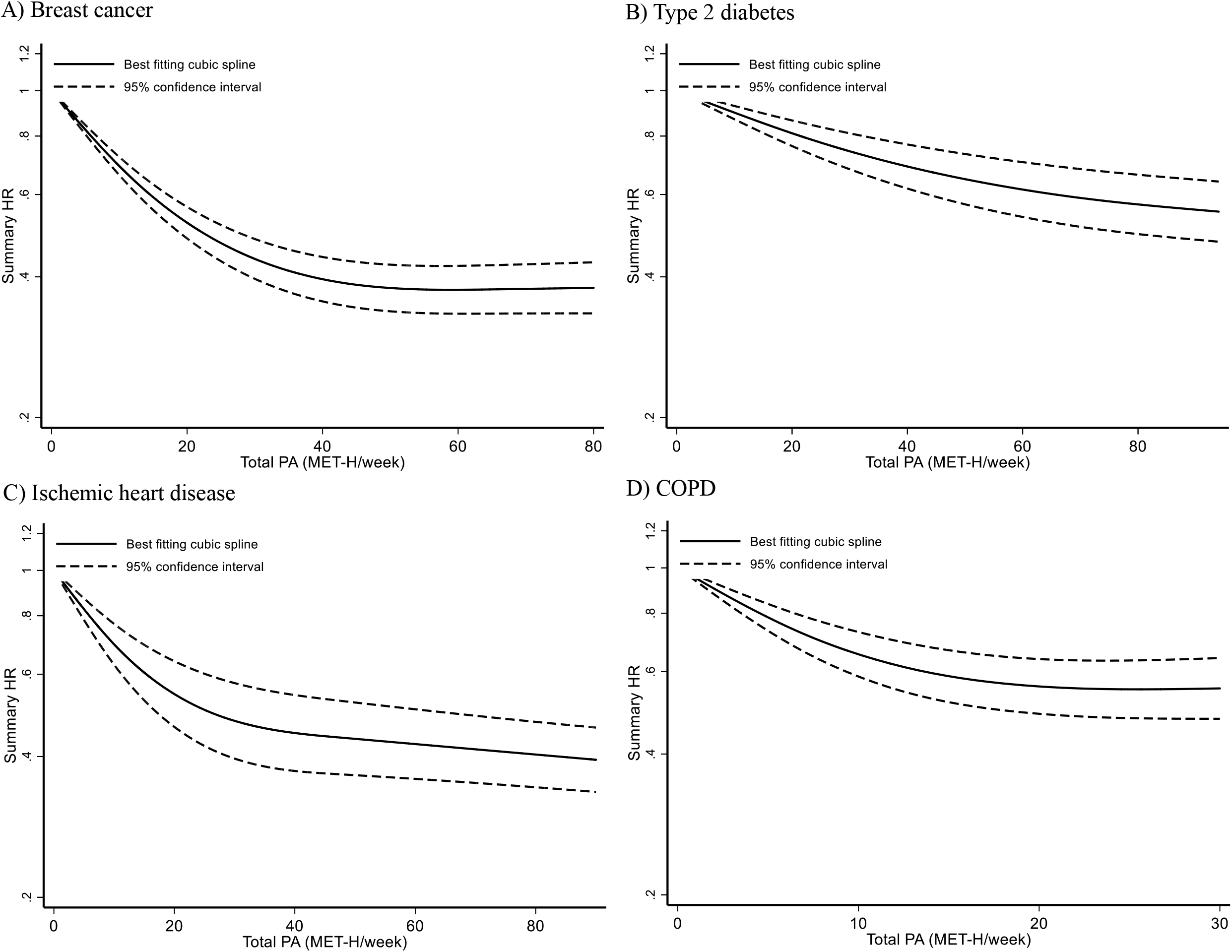
Non-linear dose–response meta-analysis for the association between post-diagnosis physical activity and all-cause mortality among adults with A) breast cancer (*n* = 12); B) T2D (*n* = 6); C) IHD (*n* = 8); and D) COPD (*n* = 2). The figure includes values up to 100 MET-h/week.

## DISCUSSION

In this systematic review and meta-analysis, higher levels of post-diagnosis physical activity were associated with a reduction in all-cause mortality in adults with breast cancer, T2D, IHD and COPD. Our dose–response meta-analysis highlights a non-linear association between physical activity levels and mortality characterized by (1) no threshold for the beneficial effect of physical activity on mortality (i.e. even low levels of physical activity are beneficial for mortality rates compared to being physically inactive), (2) a non-linear curve, where the greatest difference in mortality rates occurs among inactive compared to minimally active individuals and (3) for higher physical activity levels, the dose–response curves flatten out. The subgroup meta-analysis showed that longer follow-ups (≥10 years) lead to higher reductions of SHR. Although the effect size is higher for follow-ups that are 10 years or longer, there is an unexplained heterogeneity between the effects of physical activity within each subgroup. However, the unbalanced distribution of studies and the low overall number of studies for some subgroups make the interpretation difficult. Hence, it is uncertain whether the length of follow-up can explain the heterogeneity in the effect size. The reduction in mortality rates from physical activity were consistent and much the same after controlling for geographic areas (Asia, Europe, US, other), age (<60 years; ≥60 years), number of cases (<100, 100–500) and the risk of bias (moderate; serious). Due to a lack of studies, we were not able to determine dose–response relationships for physical activity and mortality in adults with low back pain, osteoarthritis, depressive disorder, lung cancer or stroke.

### Comparison with other studies

Our findings confirm previous linear meta-analyses, which showed a general correlation between higher physical activity levels and lower mortality rates in adults with T2D[6] and breast cancer.[7] Our linear meta-analysis reveals reductions of SHR per 10 MET-h/week that vary between the four NCDs. The lowest SHR reduction in our results was found in T2D (4%) – a somewhat lower effect than the 9.5% reduction per one MET-h/day reported by Kodama et al.[6] We found medium reductions in IHD (12%) and breast cancer (22%) and the highest SHR reductions in COPD (30%). Our applied non-linear dose–response-analysis extends and refines these previous linear analyses. The associations between different post-diagnosis physical activity levels and mortality for adults with NCDs are very similar to those recently developed for the general population.[9, 53, 54] Therefore, our results confirm the following main characteristics of the dose–response curves in the general population for adults with selected NCDs: (1) no threshold for the positive effect, (2) the most pronounced SHR reductions occurs between adults with little physical activity compared to those being physically inactive and (3) no negative effects on mortality at higher volumes of physical activity.

For higher volumes of physical activity equivalent to an energy expenditure of more than five times the weekly recommended moderate-intensity physical activity of 150 minutes and more, the dose–response curve is less clearly defined. The dose–response curves of the US Physical Activity Guidelines Advisory Committee[9] does not include physical activity levels of more than 30 MET-hours/week. Ekelund et al.[54] include higher volumes of physical activity, stating that the maximal reductions in SHR were seen at about 24 min/day of moderate to vigorous physical activity or 375 min/day of light intensity; higher volumes of physical activity are associated with a slight reduction in their benefit on mortality rates. In our study, higher physical activity levels were associated with continuously small declines in mortality rates for IHD and T2D. For breast cancer, there is a point of maximum reduction of SHR at 55 MET h/week with no additional benefits for higher physical activity levels. No data is available on higher physical activity levels for COPD.

In the total population, 70% of the maximum effect on mortality risk reduction is achieved at an energy consumption of 8.25 MET-h/week (equivalent to meeting the physical activity recommendations of 150 weekly minutes).[9] Our results indicate that in adults with NCDs, this energy consumption is associated with about 40% of the maximum achievable reduction in mortality rates. Physical activity and both overall and cardiovascular mortality after stroke were connected through a dose–response relationship where 10 MET-h/day of physical activity produced 35–46% reductions in SHR.[39] Although one study reported data on stroke patients,[39] it was not sufficient to be included in the meta-analysis. For 4 NCDs (low back pain, osteoarthritis, depressive disorder, lung cancer), we were not able to find appropriate studies for our analysis. Thus, our findings confirm the research gap in the clinical populations already identified before.[9]

### Strengths and limitations

Our study has several strengths. Its main strength is the broad and comprehensive systematic literature search for 9 NCDs that has a high relevance for public health. For the first time, our work generates a broad overview of post-diagnosis physical activity and mortality for adults with NCDs. Another strength is the applied non-linear dose–response meta-analysis that enables precise statements regarding the effective dose of physical activity for reduced mortality rates. This information helps with the adaption or development of exercise recommendations for adults with NCDs. In addition, the use of the new Robins-I tool is a methodological strength that allows for a precise estimation of the risk of bias in different domains (e.g. bias in the measurement outcome, due to missing outcome data or due to deviation from intended interventions).

Despite its strengths, this systematic review and meta-analysis has limitations that should be acknowledged. First, at the outcome level of the study, the risk of bias in the measurement of physical activity in the original studies is unknown. All studies measured physical activity levels using self-reports. Compared to device-based measurements, self-reported measures are prone to over-reporting of one’s physical activity levels.[55] Most of the studies have measured the level of physical activity only at one point in time, thus meaning that no information on changes over time is available. Moreover, different cut-off points were used by the single studies to classify the participants’ levels of physical activity. This might lower the accuracy of the dose–response curves. If the over-reporting of physical activity already plays a role at low physical activity levels, the actual high relative reduction in mortality rates of somewhat physically active persons compared to inactive persons could be underestimated. Second, at the study level, our findings are susceptible to bias derived from studies of an observational nature. Prospective observational cohort studies fail to provide conclusive evidence of a causal relationship between physical activity and mortality.[56, 57] Our results might be affected by reverse causality, as patients may tend to adjust their physical activity level according to the disease severity and prognosis. Consequently, our analysis of cohort studies does not provide a conclusive answer as to whether the reported dose–response relationships between physical activity and mortality are actually causal or only correlative. According to Hill,[58] however, our results increase the sense of confidence in a causal relationship because they display (1) a clear dose–response curve, (2) a strong association or high effect size and (3) consistent results in different studies. Third, at the review level and as reported in the study protocol,[11] we did not consider the potential differences between different physical activity intensities (i.e. light vs moderate vs vigorous), between physical activity in different contexts (e.g. leisure time physical activity vs occupational physical activity) or the interaction between physical activity and sedentary behaviour. Furthermore, our analysis is likely to be affected by small-study effects and the small number of original studies available for the sensitivity analysis.

### Implications and future research

Assuming causality, our findings have implications for adults with NCDs, physicians and other health professionals involved in physical activity promotion and exercise therapy, as well as healthcare decisionmakers and policymakers. First, our results bear importance for policymakers and those involved in public health issues. The results highlight the importance of a physically active lifestyle and support strategies to promote physical activity (e.g. the World Health Organization’s Global Action Plan on Physical Activity).[59] Second, for those creating physical activity guidelines, our findings may inform developments or updates on physical activity recommendations for adults with NCDs. Our findings reinforce low-dose physical activity recommendations that clearly demonstrate there is no minimum dose of physical activity and that effects on longevity occur at a volume of physical activity significantly below the recommended minimum dose of 150 minutes per week.[60] The “+10 minutes of physical activity per day” from Japan[61] or the “Every Step Counts” message from Germany[62] might be more feasible and efficient physical activity recommendations. Third, for physicians, our results illustrate the medical potential of exercise as medicine and encourage initiatives to anchor assess to and promotion of physical activity in routine medical care.[63] Fourth, for health professionals in the field of physical activity promotion, our results could lead to new targets for health-enhancing physical activity. Adults with NCDs are often rather physically inactive[64] and experience various barriers to physical activity, including time constraints and personal doubts about being able to participate in regular physical activity.[65–67] Completing at least 150 minutes of physical activity per week is considered by many to be overwhelming and unachievable. For adults with NCDs, non-threshold-based, low-dose physical activity recommendations could be effective while also being encouraging and easier to implement. Thus, low-dose physical activity recommendations would destroy many barriers in relation to an active lifestyle and increase the probability of success of interventions that promote physical activity.

The results also bear implications for future research. We identified a research gap: For 4 out of 9 NCDs (lung cancer, depressive disorder, lung cancer, low back pain), there were no eligible studies available. Since the associations for post-diagnosis physical activity and mortality in adults with NCDs and the total population are different,[68] future research should either conduct cohort studies on adults with NCDs or make a differentiation in the analysis of the total population between healthy people and those with an existing NCD. Furthermore, based on the considerable analyses of Ekelund et al.[54] in the overall population, future analyses for adults with NCDs should also consider different intensities and types of physical activity as well as the interaction between sedentary behaviour and physical activity. Finally, future studies should apply more reliable device-based assessments of physical activity instead of questionnaires that are prone to over-reporting.

## CONCLUSION

In conclusion, our systematic review and meta-analysis provides evidence that higher levels of physical activity are associated with lower mortality rates in adults with T2D, IHD, breast cancer, or COPD. The shape of the dose-response curves are characterized by no threshold for the beneficial effect of physical activity on mortality, and a regressive, non-linear dose-response pattern where the greatest difference in mortality rates occurs among inactive compared to minimally active individuals. There is no minimum dose of physical activity for life prolongation. Less physical activity than the recommended 150 min a week has life expectancy benefits for adults with a NCD. Our results encourage the development of low-dose physical activity recommendations for adults with NCDs.

## Data Availability

Data (including the extracted contents from the searched articles) are available upon reasonable request from Dr. Wolfgang Geidl; mail

## Footnotes

### Author’s contributions

WG had the initial idea for this review; he is the guarantor of the study. WG, SS, EM, LM and KP designed the study, including the development of the selection criteria, the risk of bias assessment strategy, the search strategy and the data extraction strategy. EM and LM conducted the bias assessment. SS conducted the meta-analysis. WG, EM and SS prepared the first draft of this manuscript. All authors contributed substantially to the drafting of the final manuscript version. All authors have read and approved the final manuscript.

## Acknowledegment

We would like to thank Anna Ryan, Lukas Janz and Katja Bartsch for supporting the process of article screening. Many thanks to PD. DR. Karim Abu-Omar for his valuable advice in the preparation of the final draft.

